# Systematic assessment of rare and *de novo* structural variants in 57 patient-parent trios using optical genome mapping

**DOI:** 10.64898/2026.01.16.26344264

**Authors:** Bart van der Sanden, Sandra Vorimo, Theresa Brunet, Aïcha Boughalem, Maureen Jacob, Ronald van Beek, Eveline Kamping, Elisa Rahikkala, Outi Kuismin, Jukka Moilanen, Katri Pylkäs, Elisabeth Graf, Sandy Loesecke, Melanie Brugger, Kevork Derderian, Ulrich Schatz, Matias Wagner, Michael Zech, Eva M.C. Schwaibold, Felix Distelmaier, Ingo Borggraefe, Katharina Vill, Lisenka E.L.M. Vissers, Juliane Winkelmann, Kornelia Neveling, Thomas Meitinger, Tuomo Mantere, Detlef Trost, Alexander Hoischen

## Abstract

Next-generation sequencing has unraveled the genetic cause for many individuals with a rare disease, but a significant number of individuals remain undiagnosed using standard of care tests. It is anticipated that structural variants (SVs) have not been fully assessed in this context. Here, we performed optical genome mapping (OGM) for 57 trios and prioritized SVs using a two-step approach. First, we systematically identified all *de novo* SVs, and subsequently we studied all rare inherited SVs. Potential pathogenic SVs were confirmed using orthogonal methods. On average, we identified 6,289 SVs >500bp per proband, primarily insertions (69.8%) and deletions (27.1%). In total, we identified 13 *de novo* SVs, confirming a *de novo* mutation rate for large SVs of 0.23 or 1 in 4-5 cases. These *de novo* SVs impacted multiple (candidate) disease-associated genes, including *NSF* and *FGF9*. Additionally, on average per sample, we identified 11 rare inherited SVs overlapping with an established OMIM disease gene or its regulatory region, including a homozygous deletion affecting *SCN9A* causing congenital indifference to pain, a maternally inherited deletion in *WWOX* causing developmental and epileptic encephalopathy, and an interchromosomal insertion in the CMTX3 locus at Xq27.1 causing X-linked Charcot-Marie-Tooth disease. In total, we identified pathogenic SVs in three individuals and candidate disease-causing SVs in five other individuals. Overall, OGM enabled the accurate detection of challenging *de novo* and rare inherited SVs. Our results suggest a potential yield of disease-associated SVs in 5-14% of index cases, demonstrating that OGM can unravel previously hidden SVs in extensively tested individuals.

## Introduction

During the past decade, the search for pathogenic variants in rare genetic diseases has involved huge efforts to sequence coding regions or the entire genome using short-read sequencing^1,2^. This approach has been highly successful, but a large part of cases still remain undiagnosed after applying standard diagnostic techniques including targeted gene-based testing, genomic microarray profiling^2^ and next generation sequencing (NGS) approaches (i.e., exome and genome sequencing)^3–5^. The results of several studies suggest that the accurate identification of (*de novo*) copy number variants (CNVs) and (*de novo*) structural variants (SVs) could still explain an important part of previously unresolved individuals with a rare disease^5–7^. For neurodevelopmental disorders (NDDs), it has been shown that *de novo* mutations (DNMs) in protein-coding genes explain the majority of the respective diseases in outbred populations^8–14^. The term DNM represents different variant types. The *de novo* rates are well established for substitutions, resulting in 44 to 82 substitution DNMs and 7 indel DNMs per generation^13,15–17^. However, they are less well-studied for SVs (including CNVs), and latest estimates suggest that the DNM rate is ∼2 orders of magnitude lower, resulting in only 0-2 *de novo* SV per generation^7,18–20^. This means that the *de novo* mutation rate per genome for especially large SVs that are not affected by tandem repeats, is much lower than the *de novo* rate for substitutions, which is suggestive for a constraint against (large) SVs^21^. Also, (*de novo*) SVs may have a more severe effect when located in non-coding and possibly regulatory regions of the genome than small variants^22^, as there is an *a priori* higher chance to disrupt a regulatory element with an SV than with a substitution or small indel. Additionally, *de novo* SVs are enriched in probands without diagnostic small variants compared to samples with a diagnostic small variant^23^. Finally, (*de novo*) SVs may have been overlooked by previous tests as these can be difficult to detect with short-read sequencing technologies and often occur mediated by repeat regions^17,24^.

Despite the substantial improvements of NGS technologies, it is often impossible to accurately map or assemble short reads from regions with SVs in highly homologous regions or repetitive sequences. Also, short-read sequencing has a bias against regions with high GC content, leading to an underrepresentation of these regions in such data sets^25,26^. Therefore, short-read sequencing fails to give a complete picture of variations in the human genome. Moreover, traditional cytogenetic techniques each have their own respective limitations. Karyotyping is limited by the low resolution (> 5 Mb in size), fluorescence *in situ* hybridization (FISH) by restricting the analysis to preselected abnormalities and genomic microarrays by the incapability to detect balanced rearrangements, such as inversions, translocations, insertions and other complex events^27^. With the recent introduction of long-read sequencing methods, the challenging regions of the genome can be better assessed^28,29^ and it is likely that long-read sequencing may allow comprehensive assessment of the full range of genomic variation with a much more complete set of SVs (latest estimates 25,000 >50bp/genome)^30^. This may be enabled by mapping (to recently improved) reference genomes, or by *de novo* assembly of a human genome. However, so far, even these most advanced sequencing technologies, when used on its own, are still not able to assess the most difficult parts of the human genome^31^, and a possible bias against SVs that are larger than actual read size may still exist^32^. Also, the additional costs, heavy computational burden, strong requirements for skilled bioinformaticians and still continuously improving bioinformatic tools, and limited read depth that is involved with long-read sequencing may still be a limitation for wider and systematic use in clinical laboratories outside a research context^20,25,28,32^.

Recently, optical genome mapping (OGM; Bionano) has proven its potential to provide a cost-effective and easy-to-use alternative for standard cytogenetic testing and next generation sequencing for the detection of chromosomal aberrations and SVs^33,34^. OGM generates images of ultra-long DNA molecules with an average N50 > 250kb and can generate up to ∼400x coverage per sample. After local consensus assembly generation, SVs such as deletions and insertions as small as 500 bp can be detected, as well as duplications, inversions and translocations when these are at least 30 kb in size. Therefore, OGM presents a much higher resolution compared to karyotyping, FISH and genomic microarrays, and it allows the detection of balanced and unbalanced events, for which *de novo* mutation rates are largely unexplored. In addition, OGM is independent of sequence context and in combination with the ultra-long molecules it enables the analysis of even the most complicated regions of the genome in contrast to DNA sequencing approaches.

In this study, we used OGM to analyze 57 trios with unexplained rare diseases including neurodevelopmental disorders (NDDs), renal anomalies, hearing loss, neuropathy and malformation. For these individuals, previous extensive genetic testing including at least exome sequencing remained inconclusive. Here, we aimed to identify *de novo* or rare inherited pathogenic SVs that had remained undetected using previous genetic testing.

## Material and Methods

### Index selection

Indexes and their parents were included by four different centers: Radboudumc (n=6), University of Oulu (n=8), Cerba (n=26) and Technical University of Munich (n=17). All indexes had a disorder of suspected monogenic origin (**Table S1**) and all but four indexes (I18, I20, I21, and I22) were born to non-consanguineous parents. For all indexes, extensive genetic testing including at least exome sequencing remained negative. For 55 of the 57 trios, varying combinations of additional genetic testing, including genomic microarray, karyotyping, short-read genome sequencing and long-read genome sequencing, was performed (**Table S1**). In none of the probands a disease-causing variant was identified prior to inclusion in this study. All participants or their legal representatives gave written informed consent. This study was approved by the Medical Review Ethics Committee Arnhem-Nijmegen under 2011/188 and 2020-7142 and the Ethics Committee of the Northern Ostrobothnia Hospital District (45/2015). Index cases from Munich were recruited within the Bavarian Genomes project (https://www.bavarian-genomes.de/ueber-uns.html) approved by the local ethics committee. Two cases (index I18 and I39) with a conclusive molecular diagnosis after this study have also been published in two separate case reports^35,36^.

### Optical genome mapping

Extraction of ultra-high molecular weight (UHMW) DNA for genome imaging was performed using SP-G2 Blood & Cell Culture DNA Isolation Kit (Bionano, San Diego, CA, USA), according to the manufacturer’s protocol. DNA labeling was performed according to the manufacturer’s guidelines using the Bionano Prep Direct Label and Stain (DLS) protocol (Bionano, San Diego, CA, USA), as previously described in Neveling et al.^34^. Labeled UHMW DNA samples were then imaged by the Saphyr instrument (Bionano). Each flow-cell was run to generate >400 Gb of data per sample using GRCh38/Hg38 as the reference (**Table S2**).

### Structural variant calling and filtering

The *de novo* assembly, variant calling and annotation were performed using Bionano Solve v3.7. The results were analyzed through a distinct *de novo* assembly pipeline as described earlier^33,34^. Identified SV calls were visualized and investigated using Bionano Access (v.1.7.2). SV calls were first compared to an OGM control dataset containing 285 control samples from apparently healthy individuals (Bionano) resulting in a prioritized list of rare SVs. These SVs were then filtered based on the different inheritance types. The filtering steps identify SVs by checking parent assemblies and molecules. For *de novo* keep SVs absent in both parent assemblies and molecules, for homozygous keep SVs present in both parent assemblies and molecules, for paternal keep SVs present only in the father’s assembly and molecules, and for maternal keep SVs present only in the mother’s assembly and molecules. Further variant analysis happened in a two-step approach. First, we analyzed the *de novo* events, and in the second step we focused on the rare inherited events.

### *De novo* SV filtering

Once filtered for *de novo* events, the SVs were visually inspected in the Bionano Access software to determine possible false positive variant calls. In our experience these false positive calls are usually enriched in alignment gaps, heterochromatic blocks or (near) centromeric repeats^37^. The remaining variants were considered high-confidence *de novo* events and subsequently separated into genic and intergenic events. For the genic events the genotype-phenotype compatibility was determined based on the proband’s phenotype and genotype-phenotype information available in OMIM. On the other hand, the intergenic events were overlapped with a list of ‘GeneHancer Regulatory Elements and Gene Interactions’ (**Table S3**)^38^. This list was downloaded from UCSC Genome Browser (https://genome.ucsc.edu/) on 20/11/2023. In addition, events were overlapped with topologically associating domain (TAD) boundaries. The TADs used to calculate these boundaries were downloaded from https://3dgenome.fsm.northwestern.edu/publications.html on 20/11/2023 (**Table S4**)^39^.

### Disease gene overlap

In order to limit the number of SVs that had to be manually analyzed after filtering for inherited SVs, we overlapped the remaining SVs with a disease gene list containing all Cancer Gene Census, G2P, MIM morbid, and Orphanet gene-disease associations (**Table S5**). The table was downloaded from Ensembl using BioMart on 20/11/2023 including the following attributes: Gene stable ID, Gene description, Chromosome, Gene start, Gene end, Gene name, Gene type, Phenotype, Source name, Study external reference.

### Rare inherited SV filtering

To increase the chance to identify relevant SVs for respective cases, we prioritized the rare inherited SVs in a two-way fashion. First, we selected all rare inherited SVs that overlapped with an extensive disease gene list (**Table S5**). For this overlap, also the first 50kb up- and downstream of the gene of the respective genes was considered as overlap to include SVs that potentially disturb regulatory elements. Second, we selected all rare SVs >500kb in size as well as all rare inversions and translocations even if they were located outside any disease gene. All rare inherited SVs in male probands were also specifically filtered for X-linked events.

### Structural variant validation

SVs that were considered to be potentially disease-causing were validated using orthogonal genetic testing, including Sanger sequencing, CNV microarray, short-read genome sequencing and targeted long-read sequencing.

## Results

All individuals included in this study remained genetically undiagnosed after exome sequencing. Some samples also previously received karyotyping, genomic microarray, short-read genome sequencing or long-read genome sequencing (**Table S1**). Reasons for the referral of these individuals (26 females and 31 males) to further genetic investigations included suspected genetic syndrome, multisystem abnormality, neurodevelopmental disorders, complex neuropathy, syndromic hearing loss, and (multiple) malformations (**Table S1**). In order to evaluate the utility of OGM for the detection of novel SVs, we performed OGM for the 57 patient-parent trios and analyzed the identified SVs.

### SV detection

OGM generated on average 690.3 Gb of data per sample across all 163 samples. The average label density was 15.3 labels/100 kb, while the map rate was 83.2% and the average N50 molecule length (≥150 kb) was 255.2 kb (**Table S2**).

On average, we identified 6,289 SVs (SD 199.3) per index sample. Of these, on average 1,702 were deletions, 4,390 insertions, 119 duplications, 77 inversions and 0.6 translocations. Of all identified SVs, on average 54.6 SVs (0.87%; SD 16.2) per sample were considered rare, meaning that these were present in less than 1% of the samples in a database comprising 285 population control samples.

### Detection of high-confidence *de novo* SVs

The occurrence of *de novo* SVs has been described as about one *de novo* SV in every six individuals^7,18^. Due to this very low per genome occurrence, which is suggestive of potential evolutionary pressure against *de novo* occurrence of large SVs, we argue that every *de novo* SV we identify is worth following up by investigating overlapping genes, regulatory elements and TADs. To identify *de novo* SVs from our OGM data, we used a specific filtering strategy (**Material and Methods**). First, we only selected rare SVs that were not identified in the assemblies and molecules of both parents. In total, this yielded 18 candidate *de novo* SVs (0.32 per individual, range 0-3). We next assessed whether these putative *de novo* SVs were true events as *de novo* events are amongst the most difficult events to detect because these resemble false positive calls. In our experience these false positive calls are usually related to issues in the reference genome, as they often occur in alignment gaps, heterochromatic blocks or (near) centromeric repeats^37^. Using the Bionano Access software, we first visualized all *de novo* SVs. Based on these visual inspections, five events were considered to be likely false positives (**Figure 1A; Table S6**). The remaining 13 *de novo* SVs consisted of five deletions, three duplications, three insertions, one inversion, and one translocation (0.23 events per individual, range 0-2) (**Figure 1B**). In total, eight of the 13 events overlapped with 20 different protein-coding genes of which five were established disease genes. The other five events were intergenic or overlapped with non-coding genes only (**Table 1**). All 13 events were then assessed regarding their pathogenicity using OMIM queries, disease specific databases and literature review. Ten *de novo* events were not considered to be (candidate) disease-causing events. Of these, four were located intergenic or affected non-coding genes that were not associated with disease. Two events were located in regions in which CNVs have been reported in healthy controls based on public databases such as the Database of Genomic Variants (DGV)^40^, and four events were located in genes that had no association with the individual’s phenotype. Evidence for these events was too limited to consider them as disease-causing candidates. The remaining 3/13 events, in three unrelated individuals, affected multiple TAD boundaries and two (novel) candidate genes, namely *NSF* and *FGF9*. These events were therefore considered to be candidate disease-causing.

**Figure 1:**
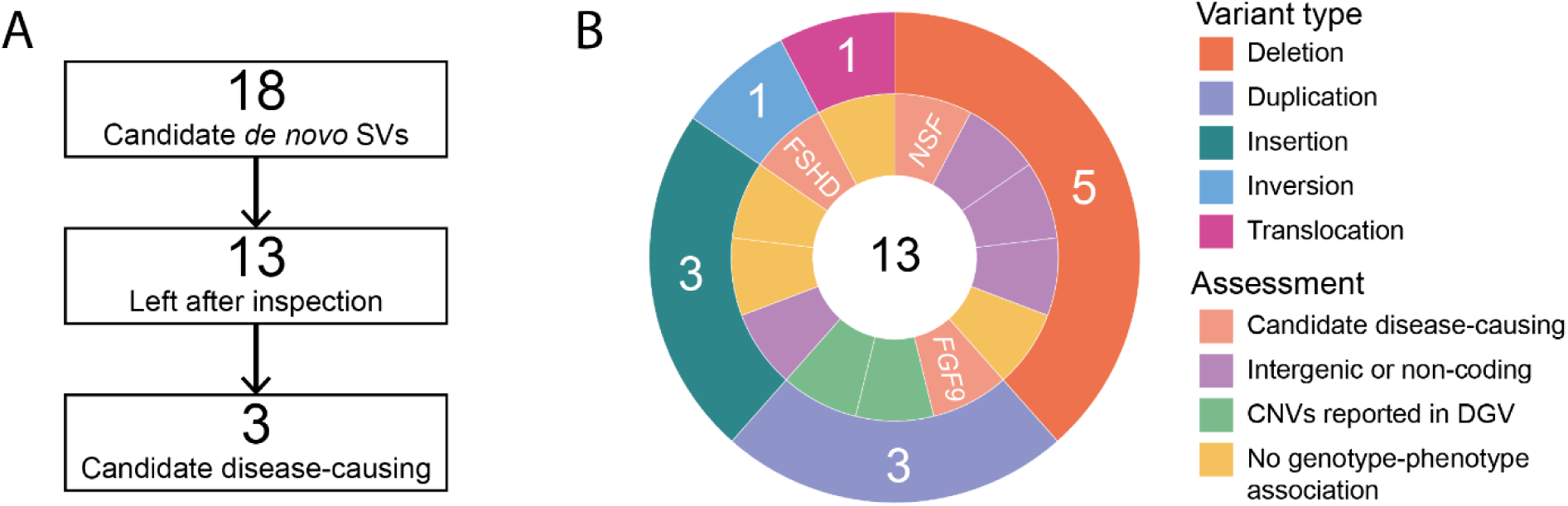
Number of identified *de novo* events. **A)** Variant interpretation flow for the *de novo* events. **B)** Variant type and assessment for each of the 13 *de novo* events left after visual inspection. Outer layer represents the variant type and the inner layer visualizes the outcome of the candidate disease-causing assessment.

**Table 1:** All 13 candidate *de novo* events remaining after visual inspection.

| Variant number | Sample ID | Genomic location | Variant type | Size (bp) | VAF | Coding/<br>Non-coding | Overlapping genes |
| --- | --- | --- | --- | --- | --- | --- | --- |
| 1 | I3 | chr17(GRCh38):g.46502601-46633474 | Deletion | 121,098 | 0.47 | Coding | <u>ARL17A</u>; <u>NSF</u> |
| 2 | I8 | chr13(GRCh38):g.56937379-57163014 | Inverted duplication | 225,636 | 0.20 | Coding | <u>PRR20A</u>; <u>PRR20C</u>; <u>PRR20B</u>; <u>PRR20D</u> |
| 3 | I19 | chr6(GRCh38):g.17516444-17768233 | Duplication | 251,790 | 0.52 | Coding | <u>CAP2</u>; <u>RPL7P26</u>; <u>SUMO2P13</u>; <u>AL138824.1</u>; <u>FAM8A1</u>; <u>NUP153</u>; <u>RNU6-190P</u>; <u>AL138724.1</u>; <u>RNA5SP204</u>; <u>Y_RNA</u>; <u>KIF13A</u> |
| 4 | I20 | chr24(GRCh38):g.9996736.4-9999647 | Deletion | 637 | 0.16 | Non-coding | <u>RBMY2QP</u> |
| 5 | I25 | chr7(GRCh38):g.144290633-144375240 | Intrachromosomal translocation | - | 0.19 | Coding | <u>AC074386.1</u>; <u>ARHGEF5</u> |
| 6 | I34 | chr4(GRCh38): g.188793071-190050588 | Inversion | 1,257,517 | 0.29 | Non-coding | <u>AC093909.2</u>; <u>LINC02508</u>; <u>AC093909.4</u> |
| 7 | I40 | chr13(GRCh38):g.21367807-21791180<br>chr13(GRCh38):g.23671562-23841571 | Interspersed<br>inverted<br>duplication<br>Duplication | 423,373<br>170,009 | 0.30<br>0.25 | Coding<br>Coding | <u>MIPEPP3</u>; <u>ZDHHC20</u>; <u>MICU2</u>; <u>FGF9</u>; <u>TNFRSF19</u>; <u>MIPEP</u> |
| 8 | I44 | chr5(GRCh38):g.20902581-21487805 | Deletion | 329,356 | 0.40 | Non-coding | <u>LINC02241</u>; <u>AC140168.1</u>; <u>AC140172.1</u>; <u>AC093274.1</u>; <u>GUSBP1</u>; <u>AC138951.1</u> |
| 9 | I47 | chr1(GRCh38):g.737536-752136 | Deletion | 1,458 | 0.47 | Non-coding | <u>AL669831.3</u>; <u>AL669831.1</u> |
| 10 | I47 | chr5(GRCh38):g.131940960-131951189 | Insertion | 1,270 | 0.23 | Coding | <u>MEIKIN</u>; <u>AC034228.4</u>; <u>AC034228.3</u>; <u>ACSL6</u> |
| 11 | I48 | chr17(GRCh38):g.47053013-47056569 | Insertion | 1,457 | 0.28 | Non-coding | <u>AC005670.3</u> |
| 12 | I51 | chr16(GRCh38):g.66482358-66495103 | Insertion | 6,040 | 0.38 | Coding | <u>BEAN1</u>; <u>AC010542.3</u> |
| 13 | I54 | chr17(GRCh38):g.57572498-57577736 | Deletion | 3,060 | 0.44 | Non-coding | <u>MSI2</u> |

Protein coding genes are underlined.

#### NSF

The SV overlapping with *NSF* in index I3 was initially classified as a duplication by the Bionano Access software. However, when validating the event using targeted long-read sequencing, the event turned out to be a 121 kb deletion (**Figure S1**). Notably, the event partially overlapped with a paternally inherited duplication in a segmental duplication region (**Figure 2A**). The patient presented with a short stature, seizures, infantile spasms and brain abnormalities. The latter two symptoms are indicative of West syndrome (**Table S1**). *De novo* pathogenic substitutions in *NSF* have a suggested dominant-negative effect and have been associated with early infantile epileptic encephalopathy^41^. Pathogenic copy number gains and losses have not been described in this region, but the pLI of *NSF* is 1.00 (gnomAD v4.1.0) suggesting intolerance to haploinsufficiency. Our finding suggests that the partial deletion disrupts *NSF* and may represent one of the first reports of *NSF* haploinsufficiency. Nonetheless, due to the complexity of the region and event, and the limited number of known patients with (*de novo*) mutations in *NSF* it is not possible to determine whether this event explains the phenotype of the patient. Further functional and clinical studies will be required to clarify the contribution of this SV to the disease pathogenesis.

**Figure 2:**
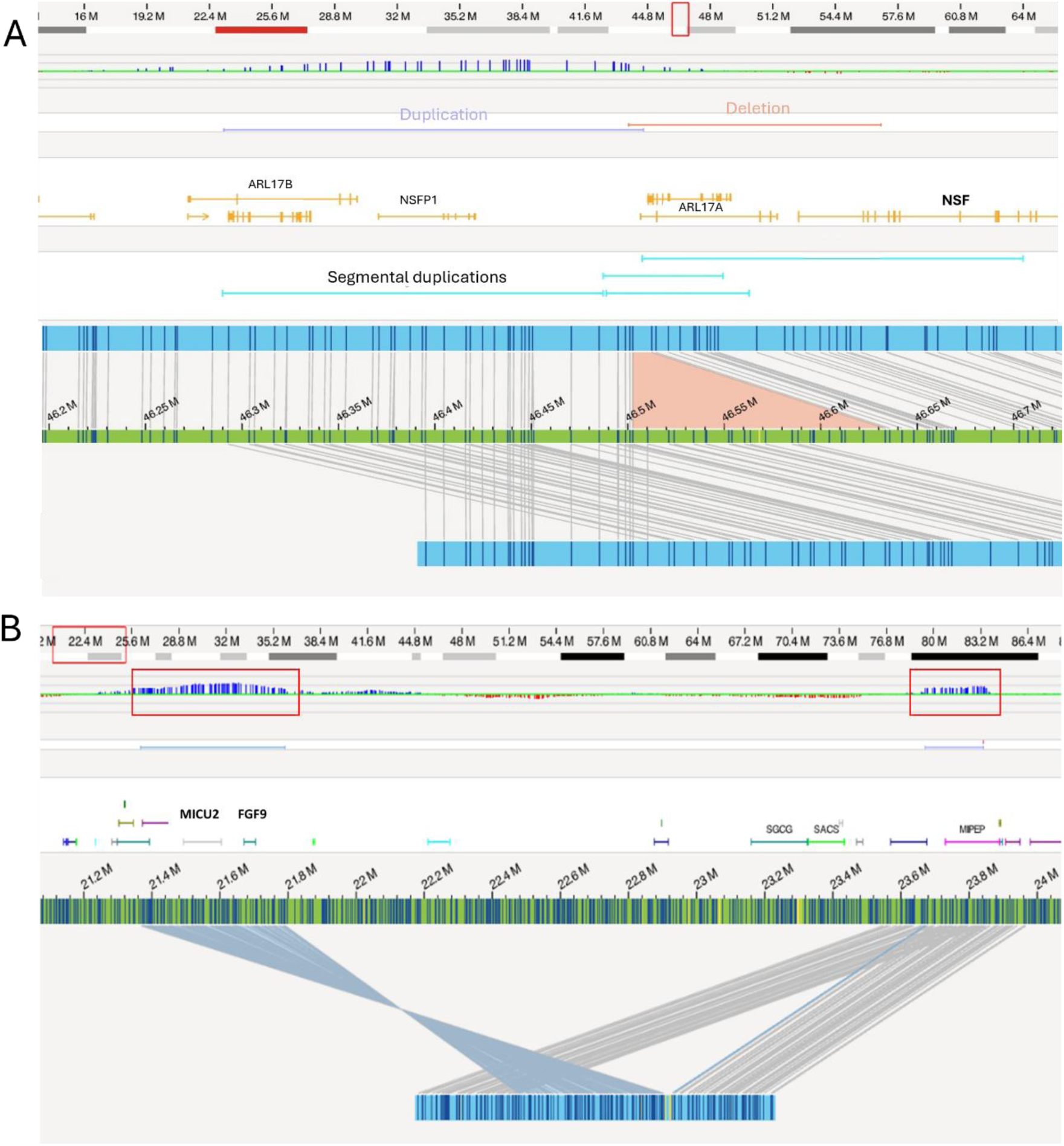
Candidate disease-causing *de novo* events. **A)** 121 kb deletion in *NSF* in index I3 (orange). This event overlapped with a paternally inherited duplication (purple) and is located in a region with many segmental duplications, which is also the proximal breakpoint region for Koolen-de Vries Syndrome. **B)** The interspersed inverted duplication involving *FGF9* in index I40. The red squares indicate CNV duplications, of which the one on the left matches the duplication detected by microarray (entailing the genes *ZDHHC20*, *MICU2* and *FGF9*). OGM analysis revealed that the duplicated region containing *FGF9* (left) is inverted and inserted ∼2 Mb downstream into the second duplicated region entailing *MIPEP*. The figure is based on an assembly that was rerun using only >300 kb molecules to provide more comprehensive information on the configuration of the rearrangement.

#### FGF9

Index I40 harbored a *de novo* event, involving an interspersed 423kb inverted duplication entailing *ZDHHC20*, *MICU2* and *FGF9* genes, which was inserted ∼2Mbs downstream into the *MIPEP* gene locus, which was also duplicated (**Figure 2B**). Originally this event was called as a single 417kb CNV gain of the *FGF9* region in microarray analysis (performed for the index only). OGM analysis now revealed higher complexity as well as the *de novo* status of the rearrangement.

Among the genes within the region, *FGF9* presents the most likely candidate involved with the patient’s phenotype. Duplications of *FGF9* are absent from gnomAD SV and DGV databases, and the gene has a high triplosensitivity score in DECIPHER (0.89). Heterozygous pathogenic *FGF9* missense variants leading to impaired FGF9 signaling have been associated with Multiple Synostoses Syndrome Type 3 (OMIM 600921)^42^. However, the patient has no typical multiple synostosis syndrome type 3 phenotype as he does not have joint fusions. Though he has a dysmorphic face, congenital atresia of the auditory ear canal and skeletal abnormality leading to hearing loss, which are phenotypic similarities with patients with *FGF9* missense variants. Notably, a recent paper has also described a patient with a *de novo* interspersed duplication rearrangement close to *FGF9* resulting in TAD disruption and *FGF9* upregulation; that patient also shared some phenotypic similarities with our patient^43^. Thus, the effect of the detected SV in our case may extend beyond simple duplication of the *FGF9* region, warranting further functional studies to demonstrate its potential pathogenicity.

### Non-coding *de novo* structural variants

The non-coding events were overlapped with a list of GeneHancer regulatory elements and other gene interactions, as well as a list of TAD boundaries. The only event overlapping with one of these lists, was the 1.26 Mb inversion in index I34 was predicted to disrupt multiple TADs. The distal breakpoint of the inversion is in the D4Z4 repeat (FSHD locus) and the proximal breakpoint does not overlap with any protein coding-gene (closest human disease associated genes being *FAT1*) (**Figure S2**). However, so far no decisive disease mechanism and genotype-phenotype correlation has been determined. Therefore, this event also remains a candidate disease-causing event.

### Inherited SVs

After filtering and analyzing the *de novo* SVs, we continued with the (rare) inherited SVs. Again, we designed a specific filtering strategy for these inherited variants. In this filtering approach, we selected rare SVs that were also identified in the assemblies and molecules of the father and/or mother of the parent-child trio (**Material and Methods**). Using this strategy we identified a total of 37.8 rare inherited SVs on average per case. To increase the chance to identify relevant SVs for respective cases, the SVs were prioritized in a two-step fashion. First, all SVs were overlapped with a list of disease genes and other regions of the genome that are associated with disease (see **Material and Methods**). This yielded on average 11.3 SVs per sample. Second, all rare SVs >500kb and all rare inversions and translocations were prioritized, which yielded an extra 0.8 SVs per sample on average. Next, we assessed the genotype-phenotype compatibility of these SVs by comparing the phenotype of the proband with the known phenotype associated with mutations in the respective gene(s) and loci. In this assessment, we also took into account whether the gene causes disease in an autosomal dominant, autosomal recessive or X-linked fashion. Hemizygous variants in male patients were also carefully assessed. In total, this yielded five candidate disease-causing SVs, which were then validated using orthogonal genetic testing. All five variants were confirmed to follow the inheritance pattern as indicated by OGM. In total, this resulted in three conclusive diagnoses and two additional variants were considered as potential disease-causing candidates.

### Conclusive diagnoses

#### SCN9A

In index I18, OGM revealed a homozygous deletion of 4.5 kb affecting the 5’UTR and first exon of *SCN9A*. Biallelic loss-of-function variants in *SCN9A* cause congenital insensitivity to pain ^44^ and hereditary sensory neuropathy type IID^45^. Index I18 also presented with this severe insensitivity to pain and an extensive case report about this patient has been published^35^.

#### WWOX

In index I29 a maternally inherited 51 kb deletion in *WWOX* affecting exon 6 was identified (**Figure 3**). Biallelic variants in *WWOX* are associated with an autosomal recessive developmental and epileptic encephalopathy^46^. As the *WWOX* locus at 16q23.1–q23.2 includes FRA16D, the second most common fragile site in the human genome, it is a hotspot for translocations and deletions and one-third of reported patients have at least one pathogenic CNV (deletions more frequent than duplications)^47^. This finding prompted a retrospective clinical re-evaluation, which confirmed a phenotype consistent with *WWOX*-related developmental and epileptic encephalopathy (DEE).

**Figure 3:**
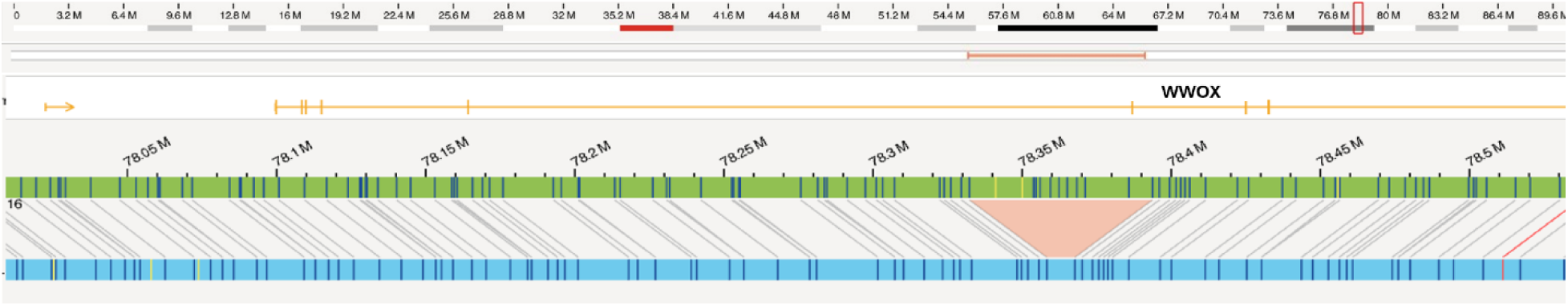
51 kb deletion in WWOX in index I29.

Subsequent reanalysis of the genome data identified a paternally inherited deep intronic variant in *WWOX* (NM_016373.4:c.107+119C>G), predicted to strengthen a cryptic splice donor site. The alternative splicing effect of the variant was later confirmed using RT-PCR. The combination of these two compound heterozygous events led to a conclusive diagnosis for this individual.

#### Xq27.1

In index I39, an individual clinically diagnosed with Dejerine-Sottas/CMTX disease, OGM analysis revealed a maternally inherited insertion from 7q31.1 into the non-coding region at Xq27.1, near *SOX3*. This region has previously been associated with CMTX3 (OMIM #302802), a rare form of Charcot-Marie-Tooth neuropathy. To date, only a single founder SV, an insertion from chromosome 8 into Xq27.1, underlying CMTX3 has been reported. In our case, OGM analysis together with manual reassessment of short-read WGS data, confirmed the molecular diagnosis of atypical CMTX3 and showed that the 122.4 kb inserted fragment contained the gene *DLD* and partial segment of *LAMB1*. Subsequent analyses confirmed that the rearrangement had arisen *de novo* in the proband’s mother. The mother had mild symptoms, such as highly arched feet, generalized muscle weakness in childhood, mild corneal clouding since birth and strabismus which was treated during childhood. This rearrangement presents a second Xq27.1 SV associated with CMTX3, but the underlying causative gene for CMTX3 still remains unknown, warranting functional follow-up studies. An extensive clinical case report about this patient has been recently published^36^.

### Candidate events

#### SLCO2A1

In index I21, we identified a maternally inherited 2.2 kb deletion in *SLCO2A1* (**Figure S3A**). Variants in this gene are associated with autosomal dominant primary hypertrophic osteoarthropathy (OMIM #167100). Both the patient and the mother presented with acne and hyperhidrosis which are overlapping phenotypic characteristics. The sister and brother of index I21 (indexes I20 and I22) both did not carry the same SV and also did not present with the same phenotypic characteristics belonging to mutations in *SLCO2A1*. However, this variant only explained a part of the full phenotype of our patient (**Table S1**). Therefore, further analysis of the genomic data of the patient and its affected sister and brother is needed to find the conclusive genetic diagnosis.

#### SEPTIN9

Optical genome mapping for index I27 identified a paternally inherited 9.2 kb deletion encompassing the 5’UTR and first exon of *SEPTIN9* (**Figure S3B**). Heterozygous variants in *SEPTIN9* have been associated with hereditary neuralgic amyotrophy leading to symptoms including muscle weakness, muscle atrophy, focal paresis, and axonal degeneration^48^. These symptoms were also present in our patient as well as the patient’s father, but still considered a candidate event because deletions around the 5’-end of *SEPTIN9* are also reported in the Database of Genomic Variant.

## Comparison with previous genetic testing

Of the three conclusive and five candidate disease-causing events, six were not identified in the initial genetic test. In addition, for index I40 only the CNVs were detected, while the full complexity of the event was missed (**Table 2**). These seven events were not identified because of the size of the event (n=2), the type of structural variation (n=2), the region in which the variant was located (n=2), or the resolution of the genetic test performed (i.e., lack of probes and coverage; n=1). The remaining event in index I29 was identified using the original test, but was not reported in absence of a second hit on the other allele. For all seven events not (fully) identified before, we went back to the standard of care (SoC) data to investigate whether knowing the approximate location of the event would help to identify the SV in retrospect. For two individuals (I34 and I39), we were able to manually infer the event identified with OGM from the original genetic testing data (**Table 2**). However, automatic variant callers did not detect these events and therefore identifying was technically impossible. Finally, for the remaining five events, no evidence was found in the SoC data. This may be due to the location of the event that was not covered by the previous test, or due to the difficult genomic location of the event in e.g., a segmental duplication region such as the deletion in *NSF* in index I3.

**Table 2:**
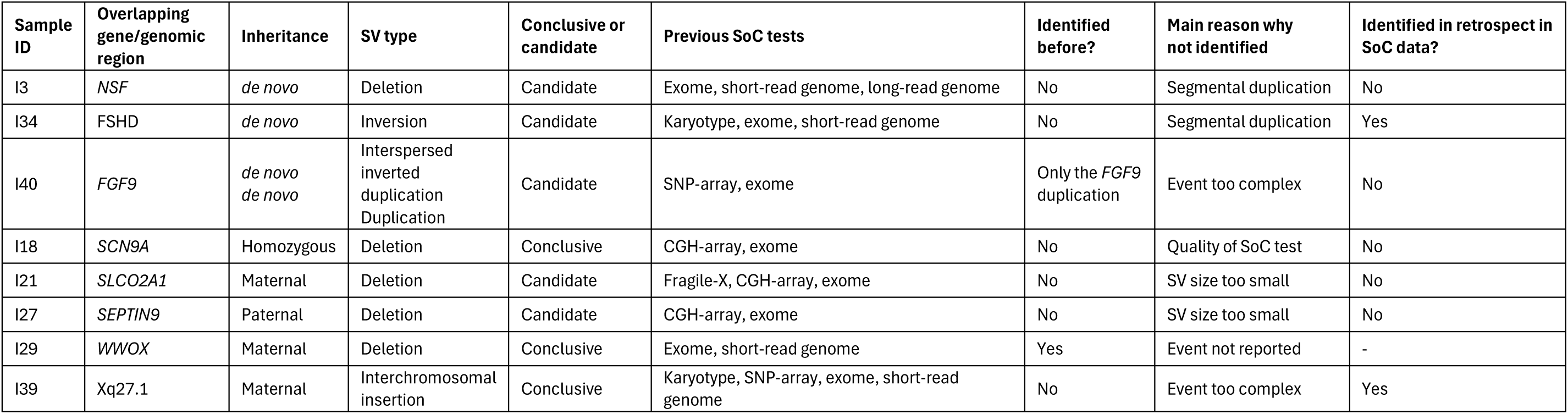
All eight candidate and conclusive disease-causing events identified in the 57 trios.

## Discussion

Here, we evaluated the utility of OGM for the detection of novel SVs by performing OGM for 57 trios with an unexplained rare disease of suspected genetic origin. This is particularly important because depending on the disease category only up to 40% of the patients can be diagnosed with the current standard of care, consisting mainly of exome sequencing and lately also short-read genome sequencing^14^. This means that many patients and their families do not receive a definitive molecular diagnosis allowing accurate genetic counselling and management strategies. It is thought that SVs make up a large part of the missing heritability of undiagnosed rare disease patients^23,32^, however the detection of SVs from short-read sequencing data is still very difficult due to read length limitations and mapping ambiguities^49^. Long-read genome sequencing (lrGS) can overcome these difficulties, but the cost of performing lrGS still limits its widespread implementation. OGM has proven to be able to accurately detect SVs >500 bp in size in even the most difficult regions of the genome and at a fraction of the cost of whole-genome lrGS. So far, OGM is mainly used to show capacity to replace SoC cytogenetic testing^33^, or in the disease causing SV identification in case reports^50,51^. Only very recently OGM has been used more systematically on cohort level^52^. Our study is amongst the first systematically studying patient-parent trios for rare inherited and *de novo* SVs.

*De novo* mutations occur during gamete formation in one of the parents or postzygotically after fertilization. These mutations have proven to play an important role in several sporadic and severe disorders^16^. *De novo* mutations are amongst the most difficult to identify, as candidate *de novo* calls may be enriched for false positive events. In addition, a recent study suggests that undiagnosed rare disease cases carry more *de novo* SVs than rare disease cases that were already diagnosed with a coding substitution or InDel variant^23^. Therefore, these *de novo* mutations can serve as an ideal proxy to assess the quality of a new genetic technology as the detection demands very high quality data in order to distinguish false positives from true *de novo* mutations. In this study, we identified a total of 18 *de novo* SVs and five of these were considered to be false positive events. These results suggest, that OGM delivers only a few false positives across 57 trios, and confirms a relatively low *de novo* SV mutation (for SVs >500bp) rate of 0.23 *de novo* SVs per genome. This number is higher than the current estimates of one *de novo* SV in every six individuals based on short-read genome sequencing efforts^7,18,23^, but lower than the *de novo* SV rate of 0.38 per genome in eight NDD trios sequenced with PacBio HiFi sequencing^20^. This may be due to the detection of SVs smaller than 500bp in size in PacBio data, which is the formal detection threshold of OGM. Or it may suggest that some of our samples harbor an undetected small variant that causes their phenotype because *de novo* SVs tend to be enriched in samples without a diagnostic small variant^23^. This low *de novo* SV mutation rate compared to the *de novo* mutation rate of e.g., substitutions, InDels and tandem repeats, suggests that there is an evolutionary constraint against *de novo* SVs in the human genome^21^. This constraint and the overall small number per genome make all *de novo* SV events interesting to follow up - especially for severe sporadic diseases such as NDDs for which the *de novo* origin of disease - causing variant is well-established^12^. In addition, the *de novo* status for SVs is part of the ACMG guidelines and therefore an important factor in pathogenicity decision. Therefore, we think that every *de novo* SV deserves thorough investigation and we first focused on these SVs. However, not all *de novo* CNVs and SVs are pathogenic *per se*^53^, and these events require careful follow-up. In our study, in total eight of the 13 identified *de novo* events overlapped with a protein coding gene. We then assessed the variant’s pathogenicity using clinical evidence, population frequency, *in silico* predictions and support from literature and databases. For two individuals, we identified an SV affecting the (novel) candidate genes *NSF* and *FGF9*. In addition, an inversion of 4q35.2 region (*de novo*) was identified in index I34. This means that we identified an interesting *de novo* SV candidate in 5.3% of the individuals. Further follow-up of these events including collecting additional pathophysiological and phenotypic evidence could upgrade these candidates to disease genes, potentially increasing the diagnostic yield. However, this often includes finding additional cases with overlapping or similar SVs, which is much more challenging for SVs than it is for small variants. This is mainly due to the very low occurrence of (*de novo*) SVs compared to small variants and because many technologies, except long-read sequencing and OGM, may miss these.

In addition to the *de novo* SVs, we also applied a different filtering protocol to identify rare inherited SVs. In fact, many of those rare SVs may have occurred *de novo* only a few generations ago^54^. In total, this filtering strategy identified five rare inherited SVs in five different individuals that were classified as likely pathogenic or a very good disease-causing candidate. The analysis of the inherited SVs resulted in a diagnostic yield of 5.3% and in another 3.4% of the individuals we identified an interesting candidate SV in X-linked or autosomal recessive disorders. These results suggest that there might be an important role for rare SVs in the missing heritability of rare disease patients and that we should continue to invest in the detection and interpretation of this variant type.

For two other patients, the use of short-read genome sequencing in parallel to OGM resulted in a conclusive molecular diagnosis. For index I44, we identified the recently described recurrent one base pair insertion in the non-coding gene *RNU4-2*^55^. For index I42, we identified compound heterozygous variants (nonsense and deep-intronic variant), in *EMC1* a gene associated with cerebellar atrophy, visual impairment and psychomotor retardation^56^. All three events were small variants and way below the detection threshold of OGM, but excluding these cases increases the yield of SVs as conclusive or candidate to 14.5% (8/55).

Even though all individuals had previously received other genetic testing, including exome sequencing, CNV microarray and some even genome sequencing, 7/8 (8%) of the likely pathogenic and candidate SVs were not (fully) identified before, which is why we also refer to these as “hidden” SVs. For the remaining SVs, previous genetic testing already indicated the maternally inherited event, but it was not reported as the event was in the recessive gene *WWOX* and a second hit was missing. Using OGM we identified the event again and subsequent analysis of a newly generated short-read genome for this individual, identified an InDel on the paternal allele. In addition, for example the duplication including *FGF9* was already called by CNV microarray, but only with OGM were we able to reveal the higher complexity of the event, demonstrating an interspersed inverted duplication as well as a second duplication event. Therefore, these results indicate that OGM can identify novel variants even in extensively tested samples, which may lead to additional diagnostic yield. For two of the “hidden” SVs we were able to detect evidence in the previously generated data in hindsight. These variants were overlooked or not prioritized in the SoC due to their quality, size, or location in the genome. Detection of SVs from exome sequencing remains challenging due to its limitation in capturing non-coding and complex regions, which results in the missed detection of most SVs. The focus on coding regions leaves larger genomic rearrangements outside the scope of exome analysis, thereby limiting sensitivity. The remaining SVs were missed because of the size of the event, the quality of the SoC test or the location of the variant in the genome. This also highlights an important benefit of performing SV detection using OGM, as the intuitive interface and availability of trio-genome filtering makes identifying hidden SVs relatively easy. Also, OGM was able to detect variants in very difficult genomic regions (e.g., in segmental duplications and telomeric regions). One example is index I3 with a 121 kb deletion affecting *NSF*. This respective genomic region is segmentally duplicated and therefore detecting the deletion using previous tests was not possible. Therefore, OGM has unique assets for rare disease studies beyond SoC^57^. Additionally, our data may still contain rare inherited SVs overlapping with genes that have not been associated with any disease before. This suggests that the yield in this cohort may still increase in the future by performing variant re-analysis.

However, OGM also has some limitations. It is not a sequencing technology and therefore does not provide sequence context and calls at breakpoint resolution. In addition, the resolution of OGM is currently at 500 bp in size, so smaller SVs are not detected by this technology, which causes that smaller disease-causing SVs are missed when only performing OGM. This means that even though it remains the highest resolution cytogenetics assay possible to date, it will never become a “single generic test” for germline testing, but will rather be used in conjunction with other tests. It is already perceivable that long-read sequencing will offer a much more complete test, and once throughput, cost and ease-of-use have caught up – this may develop towards a generic test for all variant types including DNA methylation^58^. However, the confirmation of (complex) SVs after performing specific genetic tests may still be needed and can be performed using OGM and its intuitive analysis software^59^.

In the future, the interpretation of (*de novo*) SVs detected using OGM would benefit from comprehensive SV reference databases. The number of 285 control samples used in this study provided a valuable starting point but expanding these databases with a larger and more diverse set of samples will be crucial for improving SV interpretation. This study identified several novel candidate SVs and follow-up functional studies are essential to determine their biological significance and their potential role in disease mechanisms. Evaluating rare variants, particularly *de novo* variants, requires connections with clinicians and researchers via matchmaking platforms. However, there is a lack of platforms designed for SVs in regulatory regions not directly linked to specific genes. An improved system is needed to classify and analyze these variants, enhancing the identification and functional understanding of SVs in complex genomic regions. Consequently, integrating OGM data with other techniques, such as short-read and long-read genome sequencing, will be essential for a more comprehensive analysis^50,60,61^.

Overall, we conclude that OGM resulted in a diagnostic yield of up to 14.0% of which 5.3% were already confirmed diagnoses and the remaining 8.7% were potential disease-causing candidates.

In addition, two patients were diagnosed by using short-read genome sequencing, meaning that OGM resulted in a total diagnostic yield of up to 5.5-14.5%. When considering that the diagnostic yield of most rare diseases is ∼40%, this anticipated additional diagnostic yield is of great importance and can have a major impact on the lives of the patients and their families as they now have a better understanding of their disease, the prognosis, possible treatment and the recurrence risk.

## Data Availability

All data produced in the present study are available upon reasonable request to the authors.

## Acknowledgements

TB was supported by the Kommission für Klinische Forschung (KKF), TUM School of Medicine and Health, Technical University of Munich. MZ is supported by funding from the EJP RD (EJP RD Joint Transnational Call 2022) and the German Federal Ministry of Education and Research (BMBF, Bonn, Germany), awarded to the project PreDYT (PREdictive biomarkers in DYsTonia, 01GM2302). MZ’s research is also supported by a “Schlüsselprojekt” grant from the Else Kröner-Fresenius-Stiftung (2022_EKSE.185). In addition, MZ receives funding from the Federal Ministry of Education and Research (BMBF) and the Free State of Bavaria under the Excellence Strategy of the Federal Government and the Länder, as well as by the Technical University of Munich - Institute for Advanced Study. MZ has received research support from the German Research Foundation (DFG 458949627; ZE 1213/2-1). Optical genome mapping of samples from the German project partner were funded by the Bavarian Genomes Network. TMa received funding from the research council of Finland (338374, 360442) and Sigrid Jusélius Foundation (220111). The Biocenter Oulu Sequencing Center is acknowledged for providing optical genome mapping services for the Finnish project partner. Some of the authors are a member of the European Reference Network on Rare Congenital Malformations and Rare Intellectual Disability ERN-ITHACA. ERN-ITHACA is funded by the European Union, under the grant agreement N°101156387. AH was supported by a ZonMW (The Netherlands Organization for Health Research and Development) Vici grant (No. 09150182310053). The project received funding (to KN, LELMV and AH) from the Dutch Ministry of Economic Affairs by means of a PPP Allowance made available by the Top Sector Life Sciences & Health to stimulate public-private partnerships. The aims of this study contribute to the ERDERA project (to BvdS, LELMV and AH, grant agreement N°220540), which received funding from the EU Horizon 2020 EU Horizon Europe research and innovation programs.

