## Supplementary Materials for "Systematic assessment of rare and *de novo* structural variants in 57 patient-parent trios using optical genome mapping"

[1] Department of Human Genetics, Research Institute for Medical Innovation, Radboud university medical center, Nijmegen, The Netherlands
[2] Laboratory of Cancer Genetics and Tumor Biology, Translational Medicine Research Unit, Medical Research Center and Biocenter Oulu, University of Oulu, Oulu, Finland
[3] Technical University of Munich, TUM School of Medicine and Health, Institute of Human Genetics, TUM University Hospital, Munich, Germany
[4] Bavarian Genomes Network for Rare Disorders
[5] Department of Human Genetics, Laboratoire CERBA, Frépillon, France
[6] Dr. v. Hauner Children's Hospital, Department of Pediatric Neurology and Developmental Medicine, LMU - University of Munich, Munich, Germany
[7] Department of Clinical Genetics, Medical Research Center Oulu and Research Unit of Clinical Medicine, Oulu University Hospital and University of Oulu, Oulu, Finland
[8] Northern Finland Laboratory Centre Nordlab, Oulu, Finland
[9] Institute of Neurogenomics, Helmholtz Zentrum München, Munich, Germany
[10] Institute for Advanced Study, Technical University of Munich, Garching, Germany
[11] Institute of Human Genetics, Heidelberg University, Heidelberg, Germany.
[12] Department of General Pediatrics, Neonatology, and Pediatric Cardiology, Medical Faculty and University Hospital Düsseldorf, Heinrich-Heine-University, 40225 Düsseldorf, Germany.
[13] Division of Pediatric Neurology, Developmental Medicine and Social Pediatrics, Department of Pediatrics, Dr. von Hauner Children's Hospital, Ludwig-Maximilians-University, Munich, Germany.
[14] Division of Pediatric Neurology and Developmental Medicine and LMU Center for Children with Medical Complexity, Dr. von Hauner Children's Hospital, LMU Hospital, Ludwig-Maximilians-Universität, 80337 Munich, Germany.
[15] Institute of Neurogenomics, Helmholtz Zentrum München, Munich, Germany
[16] Lehrstuhl für Neurogenetik, Technische Universität München, Munich, Germany
[17] Munich Cluster for Systems Neurology (SyNergy), Munich, Germany
[18] Department of Internal Medicine; Radboud Expertise Center for Immunodeficiency and Autoinflammation and Radboud Center for Infectious Disease (RCI), Radboud University Medical Center, Nijmegen, The Netherlands

[*]These authors contributed equally

### Figures

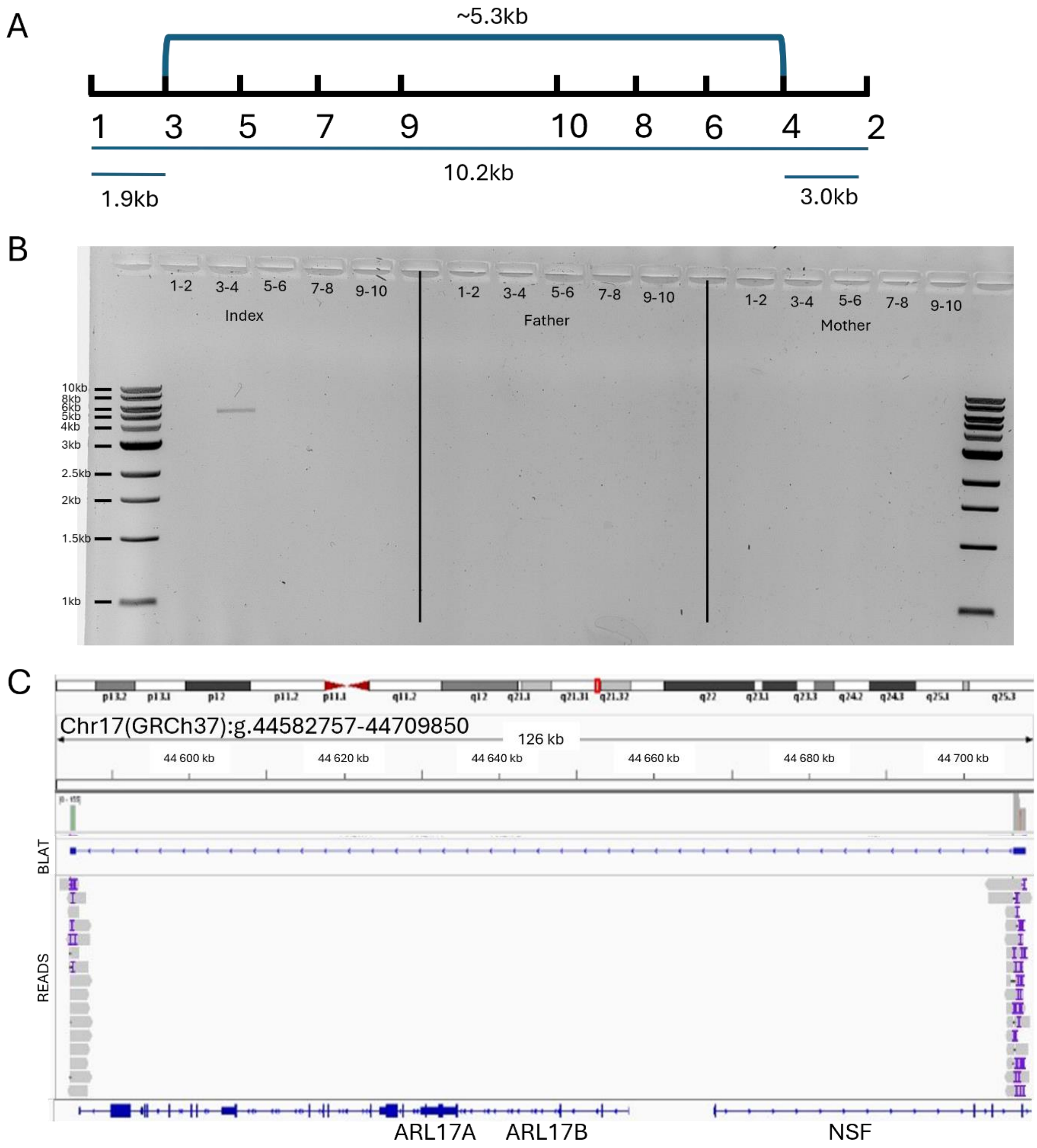


**Figure S1: Validation of *NSF* deletion in index I3.**
**A)** Using a tiling strategy, we designed five different primer pairs to validate this event. The expected size between primers 1-2 was ~10.2 kb if there was no deletion. **B)** PCR validation results after gel electrophoresis. The numbers indicate the different primer pairs. PCRs were performed for the index, father and mother. Only for primers 3-4 the PCR resulted in a product in the patient confirming a *de novo* event **C)** IGV screenshot of the long-read amplicon sequencing results validating the deletion breakpoints. BLAT analysis confirms that reads mapping to the left and right breakpoints are contiguous, consistent with a 121 kb deletion.

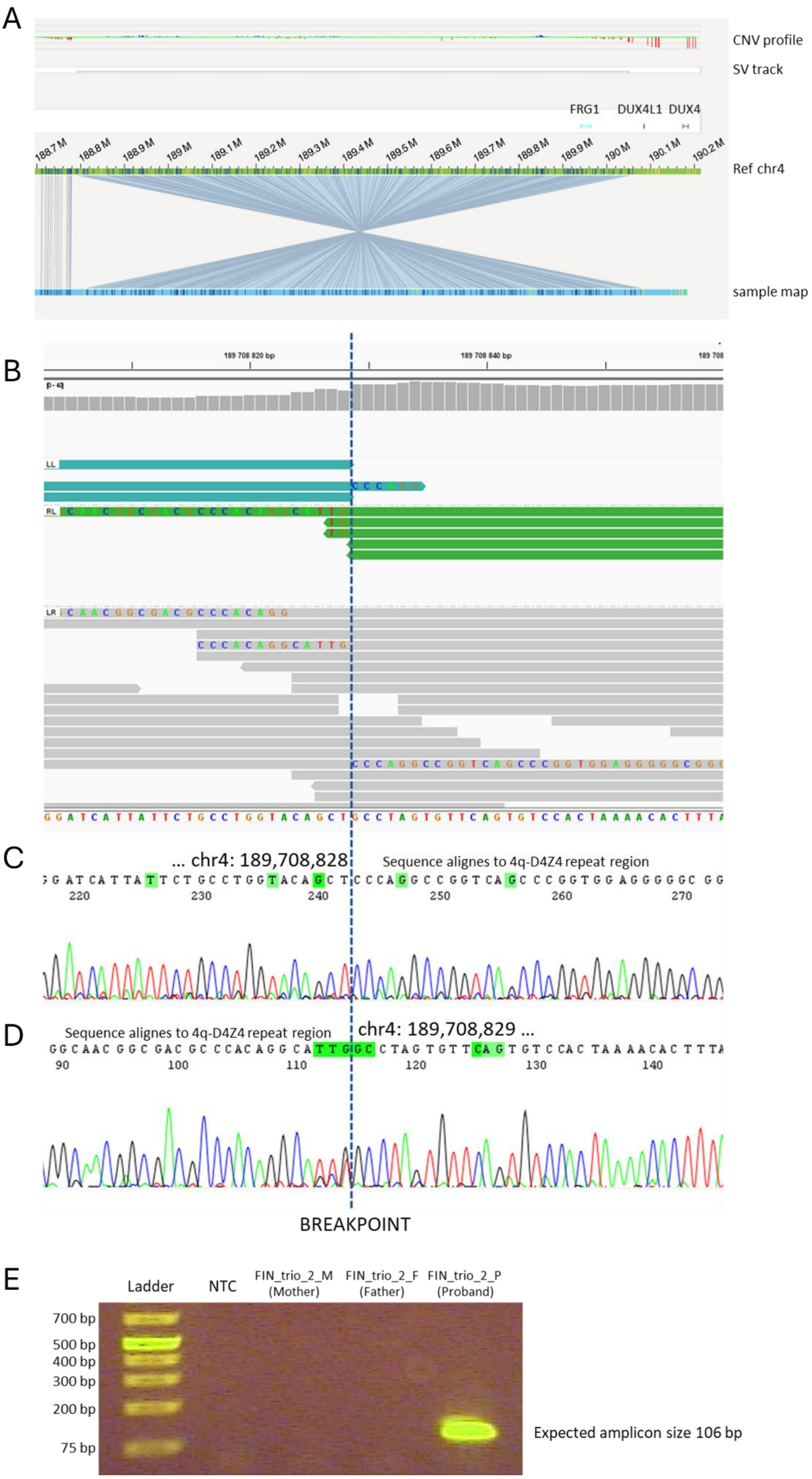


**Figure S2: Validation of subtelomeric 4q-inversion and the de novo status by short-read genome sequencing, Sanger sequencing and breakpoint spanning PCR.**

**A)** IGV view of index I34's short-read WGS data at the OGM predicted proximal 4q-inversion breakpoint (aligned to hg19). Alignments are colored by pair orientation where blue color shows mates-pairs with left-left (LL) orientation, indicating an inversion. The insert size and alignments of the LL read pairs correspond to the OGM estimated inversion size of 1.25 Mb and breakpoint locations. **B)** Sanger sequence spanning over the proximal breakpoint of the 4q-inversion at position chr4:189,708,828 (hg19) followed by sequence aligning to the 4q-D4Z4 repeat region. **C)** Sanger sequence shown in 3' to 5' direction spanning over the distal 4q-inversion breakpoint within the 4q-D4Z4 repeat region. Due to the inversion, the D4Z4 repeat region is followed by sequence aligning to position chr4:189,708,829 (hg19) onwards. **D)** 4q-inversion breakpoint spanning PCR performed for the proband and the parents confirming the de novo status of the inversion.

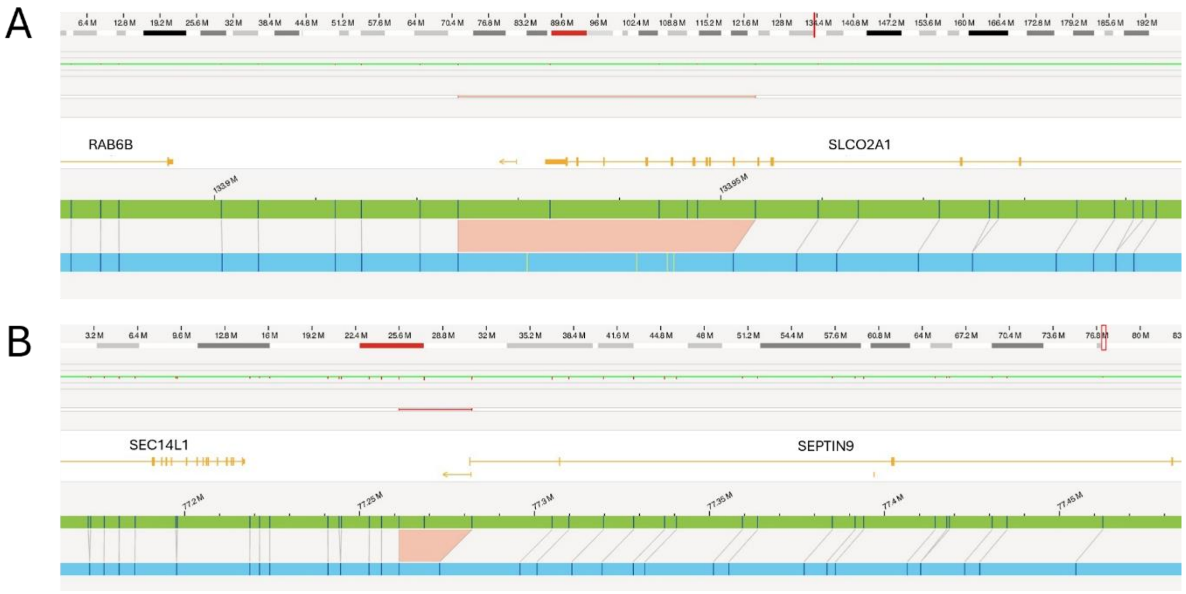


**Figure S3: Bionano Access screenshots of the two inherited candidate disease-causing events.**

**A)** 2.2 kb deletion in *SLCO2A1* in index I21. **B)** 9.2 kb deletion in *SEPTIN9* in index I27.

### Tables

**All Supplementary Tables are available in a separate Excel file.**

**Table S1: Patient overview.**

**Table S2: OGM characteristics.**

**Table S3: GeneHancer regulatory elements.**

**Table S4: Topologically Associating Domains.**

**Table S5: List of disease-associated genes.**

**Table S6: *De novo* events considered likely false positive.**
